# Barriers to cataract surgeries as perceived by visually disabled 50 years and older cataract blind participants of Nepal survey for Rapid Assessment of Avoidable Blindness

**DOI:** 10.1101/2024.10.12.24315381

**Authors:** Ranjan Shah, Sailesh Kumar Mishra, Rajiv Khandekar, Parikshit Gogate, Yuddha Dhoj Sapkota, Reeta Gurung, Mohan Krishna Shrestha, Islay Mactaggart, Ian McCormick, Brish Bahadur Shahi, Matthew Burton

**Affiliations:** Nepal Netra Jyoti Sangh, Kathmandu, Nepal; Department of Ophthalmology & Visual Sciences, Faculty of Medicine, The University of British Columbia, Vancouver, Canada; Community Eye Care Foundation, Dr. Gogate’s Eye Clinic, Pune, India; Department of Ophthalmology, D.Y. Patil Medical College, Pimpri, Pune, India; School of Health Sciences, Queens University, Belfast, UK; International Agency for Prevention of Blindness, South East Asia office, Kathmandu, Nepal; Nepal Eye Program, Tilganga Institute of Ophthalmology, Kathmandu, Nepal; International Centre for Eye Health, London School of Hygiene and Tropical Medicine, London, UK; Ministry of Social Development, Karnali Province, Nepal

## Abstract

**Purpose:** To identify the main barriers and determinants to cataract surgery as perceived by 50 years and older Nepali people with severe visual impairment & blind due to cataracts.

**Methods:** This was part of the Rapid Assessment for Avoidable Blindness (RAAB), held in all provinces of Nepal from 2018 to 2021. Cataract blindness was defined as a person having the best-corrected vision, <6/60 in the better eye, and an unoperated cataract, which was the principal cause of visual disability. The participants were interviewed using a pretested questionnaire with seven known barriers. The demographic information was correlated with the barrier score.

**Results:** We surveyed 718 cataract blinds. Two-thirds of the participants were females. Four in ten were aged 50 to 59 years. The main barriers perceived were ’need is not felt’ (237; 33%), cost associated with surgery (218; 30%), lack of access (93; 13%), fear of surgery (88; 12%), nobody to accompany (40; 6%), unaware of surgery (18; 3%), and treatment denied (24; 3%). The barriers were not significantly different in females than in males. (P = 0.85). The provincial variations of barriers were significant. (P <0.001). High cost was a perceived barrier in all provinces except Gandaki. Access to treatment was a barrier in the Gandaki province (38%). One in four participants in the Madhesh and Bagmati provinces feared surgery. Nearly half of the cataract blind in the Madhesh and Lumbini province did not feel ‘need for restoring vision’.

**Conclusions:** To improve cataract surgery uptake, identified barriers, like lack of awareness, low visual need, and high cost must be addressed. The strategies could be devised according to provincial barriers but similar to both genders and all 50 years and older cataract blind. Offering low-cost cataract surgery, financial assistance and health promotion to improve awareness and remove fear were recommended.

## INTRODUCTION

Cataract surgery is the 2^nd^ most cost-effective public health initiative to address avoidable blindness. [1] Therefore, the World Health Organization and professional agencies supported cataract surgery provisions to address avoidable visual impairment due to cataracts. [2] All efforts were focused to identify and address the factors affecting the uptake of cataract surgeries, especially in low and middle-income countries.[3] The proposed strategies included awareness campaigns, using success stories, reducing direct and indirect costs, reaching the unreachable, and maintaining and monitoring the quality of cataract surgeries. [4] To identify the magnitude and underlying causes of avoidable blindness, mainly unoperated cataracts, various countries undertook Rapid Assessments for Avoidable Blindness (RAAB) in the last three decades.[5] The findings helped the national and subnational administrators plan better the prevention of blindness programs and eye care services. Identification of barriers to cataract blindness was also part of such initiatives. [5] Few studies in Nepal reported barriers to cataract surgery, which resulted in low uptake. They included high cost, fear of surgery, distances from eye care services, and lack of awareness. [6, 7, 8] Female gender remained a significant barrier to access to cataract surgeries in countries of Southeast Asia, including Nepal. [9]

To the best of our knowledge, at the national level, including all provinces of Nepal, RAAB and barriers for cataract surgeries have not been identified in the last few years to study the impact of national and international initiatives to strengthen eye care and reduce avoidable blindness.

We present the main barriers and determinants to cataract surgery as perceived by 50 years and older Nepali people with severe visual impairment due to cataracts.

## METHODS

The survey adhered to the principles outlined in the Declaration of Helsinki. All eligible participants were informed about the survey’s purpose and procedures, and written informed consent was obtained prior to their enrolment. This process ensured voluntary participation in both data collection and examination procedures. Additionally, appropriate remedial actions were taken to address any eye or other health-related issues identified among participants.

Ethics approval for the survey was also obtained from the Ethical Review Board of NHRC, a national regulatory body for health research of Nepal, under the Ministry of Health, Government of Nepal. A letter from the Department of Health Services (DOHS) was also circulated to local government authorities to ensure necessary cooperation for the survey team and data collection.

During the RAAB survey in Nepal, we identified 920 persons with severe visual impairment. Of them, 84% (775) were due to unoperated cataracts or complications of cataract surgeries. We approached all cataract blinds to participate in the barrier study. Data collection was conducted door-to-door by trained teams led by ophthalmologists in each selected cluster. Participants aged 50 years or older were identified using citizenship cards or historical events to verify age. Eligible participants were those who had resided in the cluster for at least six months. Visual acuity assessments, anterior segment exams, and fundus evaluations were performed using standardized protocols. In some provinces, blood glucose tests were conducted to screen for diabetic retinopathy. Data were recorded using the mRAAB application on tablets and synchronized to a central server after review by a supervising ophthalmologist. Operational definition: Blindness was defined as a person having the best-corrected vision, <6/60 in the better eye.

To ensure accuracy, all teams underwent standardized training by certified RAAB trainers, with practical field exercises. A minimum inter-observer agreement (Kappa score of 0.6) was required for key assessments. The RAAB methodology, endorsed by the ICEH and WHO, was used, and data were synchronized in real-time to reduce errors. Cluster random sampling and standardized enumeration ensured representative sampling, while RAAB software facilitated consistent sample size calculations and data management across provinces.

The data collection for Province 01 (Koshi)-started on 05 June 2019 to September 2019, for Province 02 (Madhesh) from 18 December 2019 to 18 December 2020, for Province 03 (Bagmati) from 12 June 2019 to December 2019, Province 04 (Gandaki) from 05 June 2029 to October 2019, for Province 05 (Lumbini) from 03 October 2018 to December 2018 and for Province 06 (Karnali)-03 January 2019 to 02 January 2020 Province 07 (Far western) from 19 July 2020 to 19 July 2021

### Statistical analysis

We transferred the barrier-related survey data from the master file of the RAAB study after delinking from other survey information. The spreadsheet of the statistical package for Social studies was used for univariate analysis using the parametric method. The qualitative variables were presented as frequency and percentage proportions. The normally distributed numerical variables were presented as mean and standard deviation. To compare the outcomes of two subgroups, we used students’ T-test and estimated Risk ratio, 95% confidence interval, and two-sided P value. For more than two subgroup comparisons, we presented the chi-square value, degree of freedom and two-sided P value. A P-value of <0.05 was considered statistically significant.

## RESULTS

We interviewed 718 of 760 Nepalese aged 50 and older with severe visual impairment and blindness due to cataracts. Their demographic profile is given in **Table 1**. Nearly two-thirds of participants with blinding cataracts were females. Four in ten cataract blinds in this study were 50 to 59 years of age. Lumbini contributed one-third of the surveyed participants, while Madhesh contributed one-fourth of the cataract-blind participants, both in the relatively densely populated plain (terai) region.

**Table 1.**
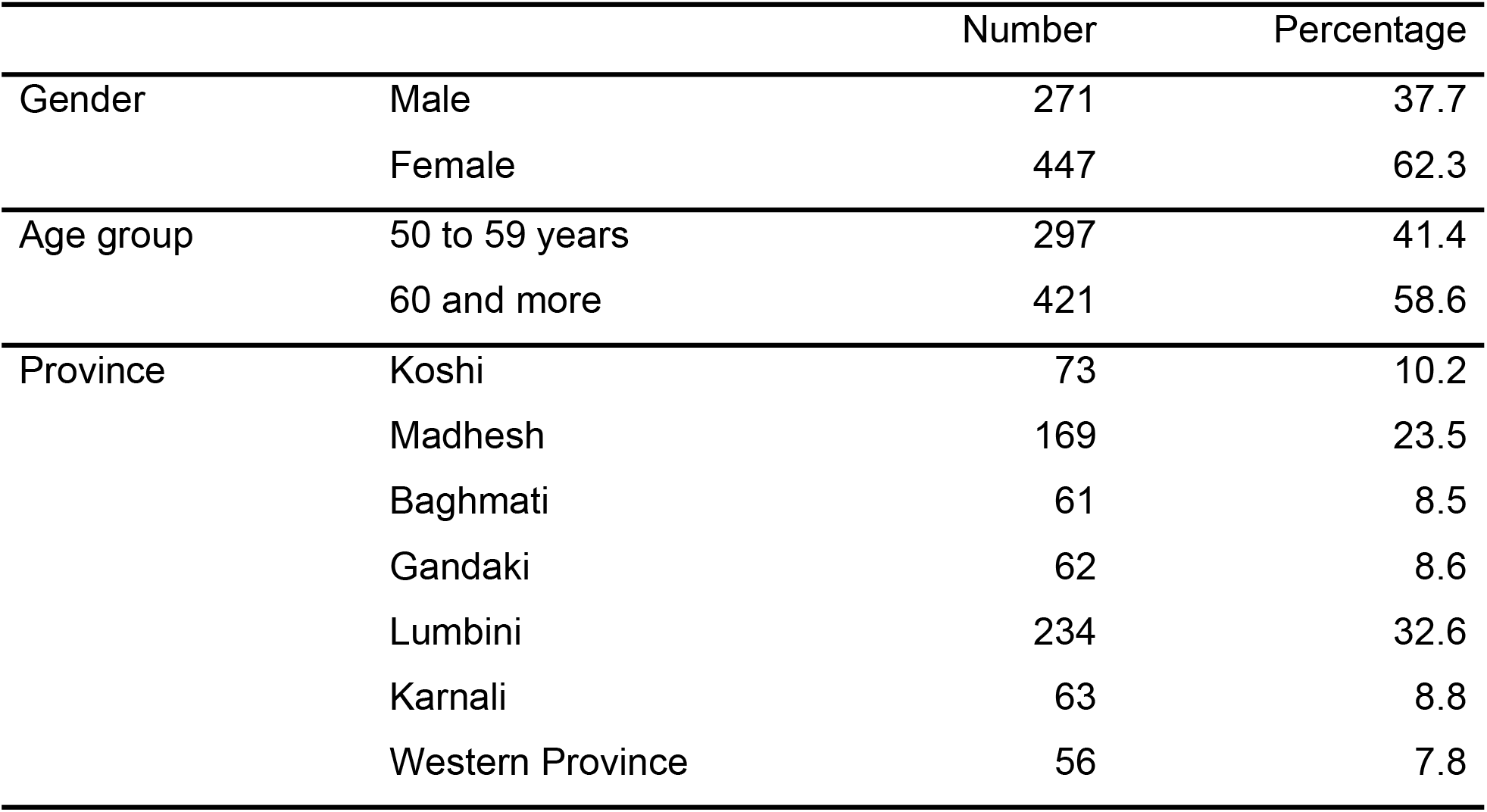
Demographic profile 50 years and older cataract blind surveyed for the barrier to cataract surgery in Nepal.

**Figure 1** shows the proportion of principal barriers among surveyed cataract blinds. According to nearly one-third of cataract blinds, less visual needs and the high cost of cataract surgeries were the main barriers to cataract surgery. One in eight surveyed participants felt fear of cataract surgery and lack of easy access to cataract surgery services.

**Figure: 1.**
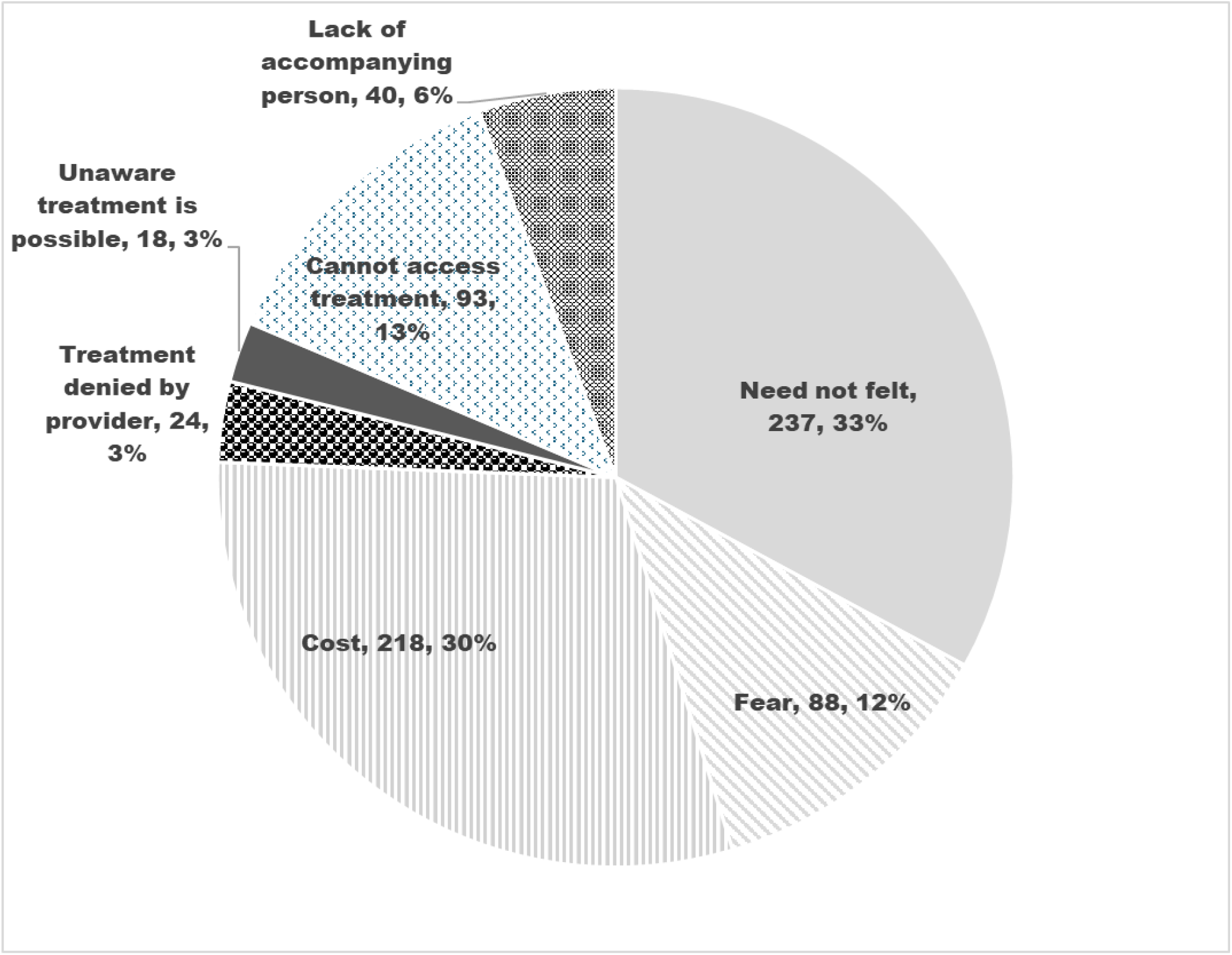
Principal barrier that cataract blind participants faced to avail of cataract surgeries Upper chart one is more suitable to avoid cost of color printing to authors.

**Table 2** shows barriers to cataract surgery in male and female cataract blind individuals. The barriers perceived by females were not significantly different from those perceived by males. High cost and fewer visual needs affected one in three participants of both genders equally.

**Table 2.** Barriers to cataract surgery in Nepal by gender.

|  | Male |  | Female |  |
| --- | --- | --- | --- | --- |
|  | Number | Percentage | Number | Percentage |
| Felt no need | 92 | 33.9 | 145 | 32.4 |
| Fear | 28 | 10.3 | 60 | 13.4 |
| Cost | 87 | 32.1 | 131 | 29.3 |
| Treatment Denied | 6 | 2.2 | 18 | 4.0 |
| Unaware of treatment possible | 9 | 3.3 | 9 | 2.0 |
| Cannot access treatment | 36 | 13.3 | 57 | 12.8 |
| Local reasons | 13 | 4.8 | 27 | 6.0 |
| Total | 271 | 100 | 447 | 100 |

Chi square = 0.1, df = 13, P = 0.85

**Table 3** shows the percentage proportions of different barriers perceived by cataract blinds in various provinces of Nepal. In the Koshi province, high cost and local reasons were the main barriers. In the Madhesh province, fear of surgery, low visual needs, and high cost of cataract surgeries were the main barriers. In the Baghmati province, high cost and fear of surgery were the main contributors to barriers. Low visual needs were the only significant barrier to cataract surgery in the Gandaki province. Lumbini province, with the highest numbers of cataract blind participants, showed high cost and low visual needs as the main deterrent for cataract surgery. The participants of Karnali provinces expressed that a lack of easy access to cataract surgeries and the high cost of services are leading barriers. In the far western province, Sudur Paschim, nearly half of the cataract-blind participants faced high costs as the main barrier.

**Table 3.** Barriers as perceived by cataract blind 50 plus Nepalese by province.

Chi square = 0.1, df = 13, P = 0.85 Table: 3 Barriers as perceived by cataract blind 50 plus Nepalese by province
|  | Koshi<br>(N = 56) | Madhesh<br>(N = 169) | Baghmati<br>(N = 61) | Gandaki<br>(N = 62) | Lumbini<br>(N = 234) | Karnali<br>(N = 63) | Western<br>Province<br>(N = 56) |
| --- | --- | --- | --- | --- | --- | --- | --- |
| no need felt | 21.9 | 49.7 | 16.4 | 27.4 | 41.0 | 6.3 | 17.9 |
| Fear | 9.6 | 24.3 | 27.9 | 8.1 | 2.6 | 1.6 | 19.6 |
| Cost | 28.8 | 21.9 | 21.3 | 14.5 | 35.9 | 39.7 | 51.8 |
| Treatment Denied | 2.7 | 4.1 | 8.2 | 11.3 | 0.0 | 3.2 | 1.8 |
| Unaware of treatment possible | 4.1 | 0.0 | 6.6 | 8.1 | 0.9 | 1.6 | 5.4 |
| Cannot access treatment | 9.6 | 0.0 | 9.8 | 12.9 | 19.7 | 38.1 | 3.6 |
| Local reasons | 23.3 | 0.0 | 9.8 | 17.7 | 0.0 | 9.5 | 0.0 |
| Tota | 100% | 100% | 100% | 100% | 100% | 100% | 100% |

Chi square = 232, df = 36, P <0.001.

**Table 4** shows barriers to cataract surgeries in Nepal in different studies.

**Table 4.**
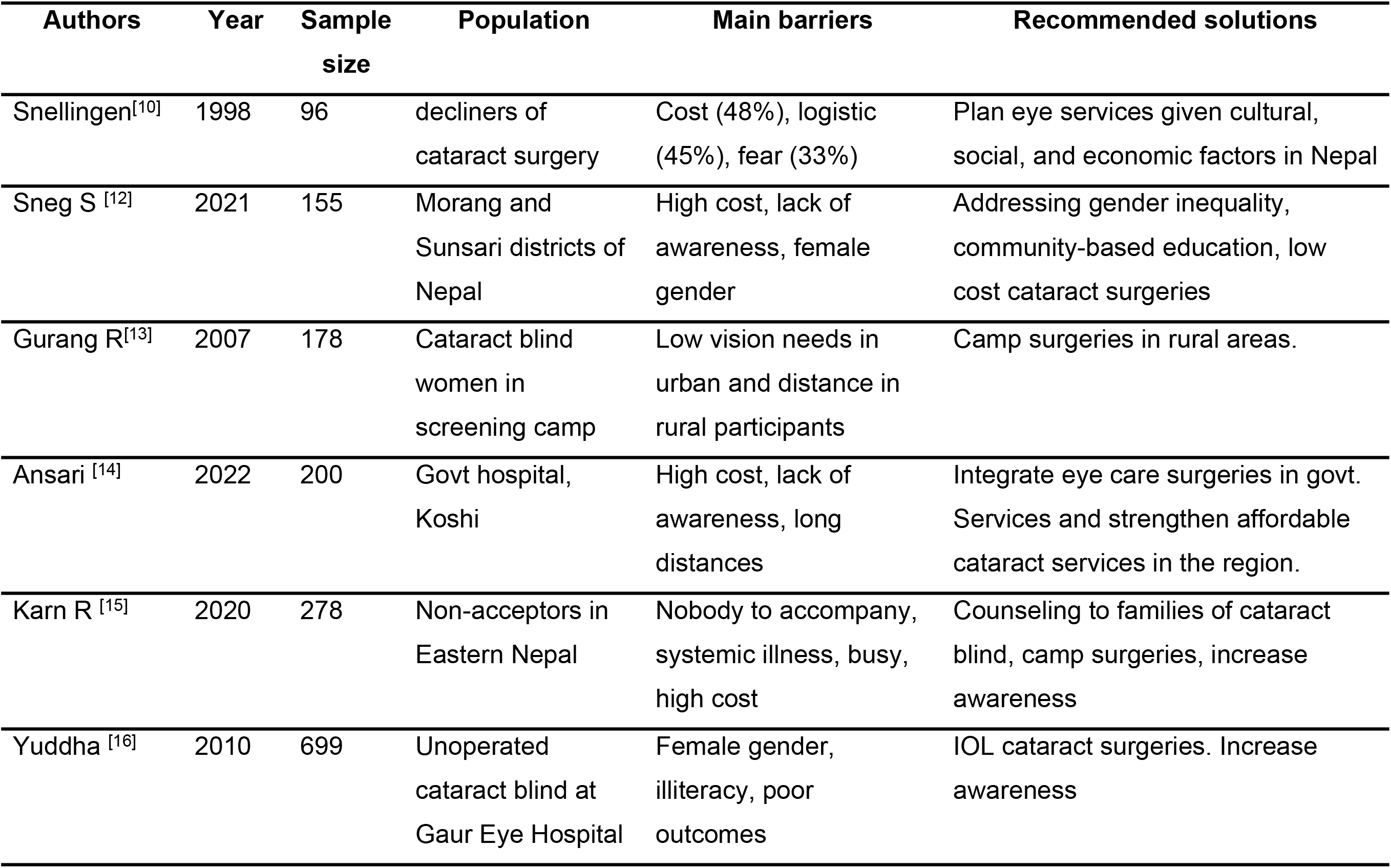
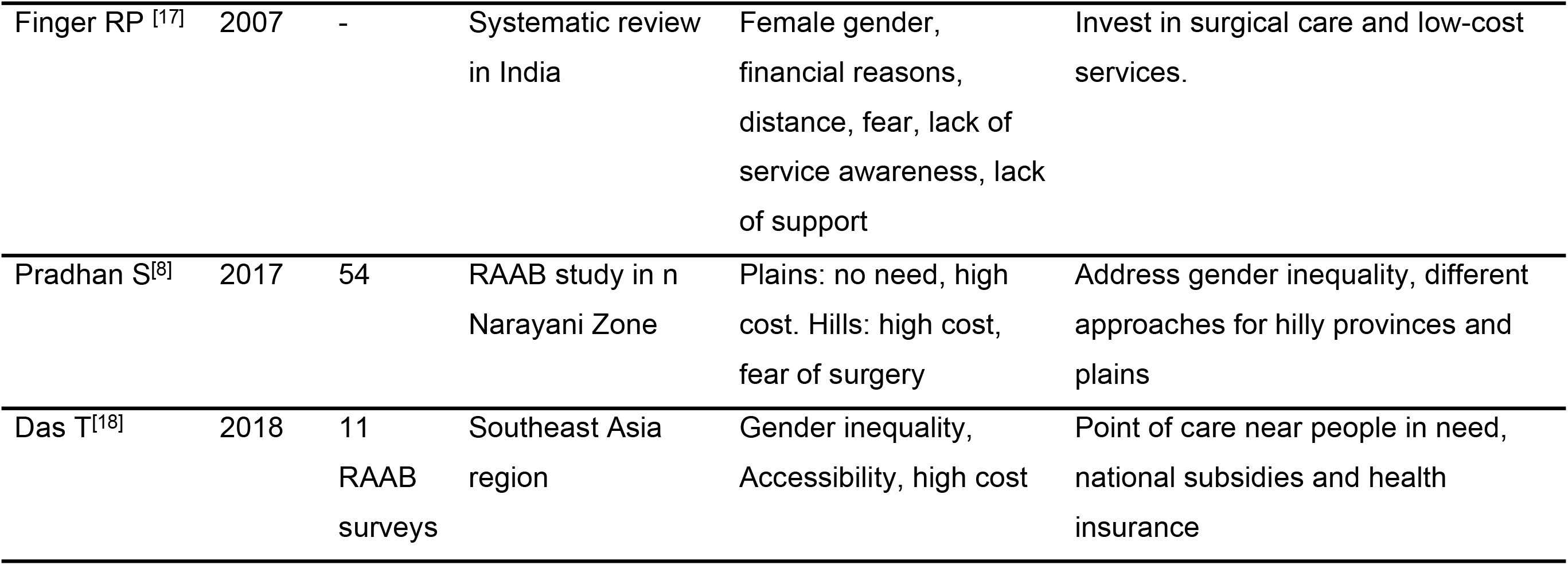
Barriers to cataract surgeries in Nepal in different studies.

## DISCUSSION

Nearly all identified Nepali persons aged 50 years and older with severe visual impairment due to cataracts identified in the RAAB survey provided information on perceived barriers to cataract surgery. With two-thirds of them being females, female gender seems to be a significant barrier. No felt need, high cost associated with surgery, fear of surgery, lack of awareness and lack of access to treatment were the present study’s main barriers. The provinceal variation was significant. Lumbini and Madesh provinces of Nepal had high cataract blinds due to a lack of felt need, fear and perceived high cost associated with cataract surgery. In the Karnali province, over one-third of the cataract blind perceived high costs and lack of access to the services as the main barriers. In the Bagmati province, one in ten cataract blinds were denied treatment by existing eye care services. This may have been due to other morbidity. As part of the RAAB survey in all provinces of Nepal, we found that cataract-related visual impairment constituted a significant contributor. Although rapid progress is being made in strengthening eye care services by government and non-governmental organizations, the female gender and high cost are still substantial barriers. The differential barriers in the provinces found in this study could help provinceal prevention of blindness units adopt national policies with variations to address these barriers. Since the present study was community-based, it is less likely to be influenced by social desirability bias than a hospital-based study. They also cover those not reaching the hospital, giving an accurate picture of barriers among those not reaching the hospital.

In the present study, lack of felt need, high cost and gender inequity among cataract blinds were the foremost barriers. This was also found in other studies held in different provinces of Nepal during outreach screening campaigns and hospital-based studies. [10,11-16] They matched with barriers documents in other low socio-economy countries in Asia, Africa, and Latin America. [17-19] The review of the impact of VISON 2020 in Nepal has revealed that NGOs and the private sector mainly provide cataract surgery services and are costly for the visually disabled, and insurance-based service provision is failing due to dropouts of premium payments. Population growth, aging, inequitable distribution of resources, and lack of integration between levels of eye health are a few challenges to reaching the goals of universal eye health and VISION 2030. [20] The proportion of females amongst the cataract blind individuals was two times more than that of males. However, the barriers perceived by females were not significantly different from those perceived by males. This matches the findings of Ye et al., who reviewed gender-specific barriers in countries of South Asia. [21] Khanna et al. attributed gender inequity to gender-defined social roles, low literacy, and urban-rural differences. [22] Nepal is ranked 166 on the Global Gender Gap Index (GGGI) with a 0.664 score. [23] Thus, gender inequity in cataract blinds may be due to differential access and priorities for cataract surgeries among males. The perceived proportion of high cost and lack of awareness and access to treatment were similar in females and males. Special emphasis on reducing the cost and awareness among females and making service areas women-friendly are some of the time-tested strategies for addressing this barrier and thereby increasing the uptake of cataract surgeries. [24] The peripheral eye camps and subsidized and free surgery offered by service providers in Nepal is effort towards this.

Four of the ten cataract blinds in our study were 50 to 59 years of age. We could not study the differential perceived barriers by age groups. However, this group with a longer life span and contribution to the economy must be addressed as a priority. A study of 60 years and older cataract blind in Sri Lanka noted high cost, lack of awareness, and other family issues as barriers to cataract surgeries. [25] We noted provinceal variation in perceived barriers to cataract surgeries in Nepal. Provinces like Karnali and western provinces with low GDP had high costs associated with cataract surgery as the major barrier. [26] In Bangladesh, Kenya, and the Philippines, perceived barriers to cataract surgeries differed. [27] It will be interesting to associate the barriers noted in different provinces of Nepal with eye care service distribution, terrains, and population density.

Health literacy campaigns can address the lack of awareness and fear of surgery. They could be gender sensitive and use successfully operated cases as champions to motivate prospective cataract blind, [4, 28] as lack of felt need was the commonest barrier.

Reducing the direct and indirect costs has been shown to increase cataract surgery uptake. This is possible by screening camps in rural areas, transporting identified cataract blinds from homes to surgery centers, providing them and caretaker gender-specific services, and reducing the cost of cataract surgeries through insurance, government subsidies, and incentives to ophthalmologists in the private sector. [4, 12, 29,30] Even if the surgery is free, there are costs in terms of travel, food, loss of wages for the patient and attendant. A randomized trail from Vietnam found that helping for other out of pocket expenses of cataract surgery was very helpful. [31] Information on past cataract surgery in these cataract blinds was also unavailable, so we could not study perceived barriers affected by the quality of previous cataract surgeries. Among people with cataracts and visual disability, we could not further divide them with SVI and with vision <3/60 in the better eye. Therefore, barriers in comparison with other studies in countries where barriers were evaluated based on vision <3/60 should be done with caution. The sample of cataract blinds in provinces may not represent all cataract blinds, and therefore barriers at the subgroup level provide trends only.

It will be interesting to associate perceived barriers noted in the present study with cataract surgery rates and effective cataract surgery rates in different provinces of Nepal.

## Conclusions

Universal efforts to address avoidable blindness will make inroads only if those suffering from cataracts and visual impairment are correctly understood. The barriers found in studies that review the perceived obstacles of those reaching and not reaching hospitals provide good insight into health planners and service providers.

The Nepal Netra Jyoti Sangh and its affiliated hospitals provide free and subsidized surgery to the populace. They conduct regular eye screening in peripheral centres and do surgical eye camps in secondary hospitals to reduce the barriers of cost and distance. Ophthalmologists and other eye care providers should ensure that patients who turn up for cataract surgery are operated upon, even with ocular co-morbidities and systemic conditions, as they may not come for treatment again soon. ‘Lack of felt need’ was the commonest barrier. Further research is needed to understand why persons with visually impairing cataract were not taking up surgery in spite of relatively economical, safe surgery.

## Data Availability

The data is available in the RAAB repository. Nepal Netra Jyoti Sangh is responsible for the data

---

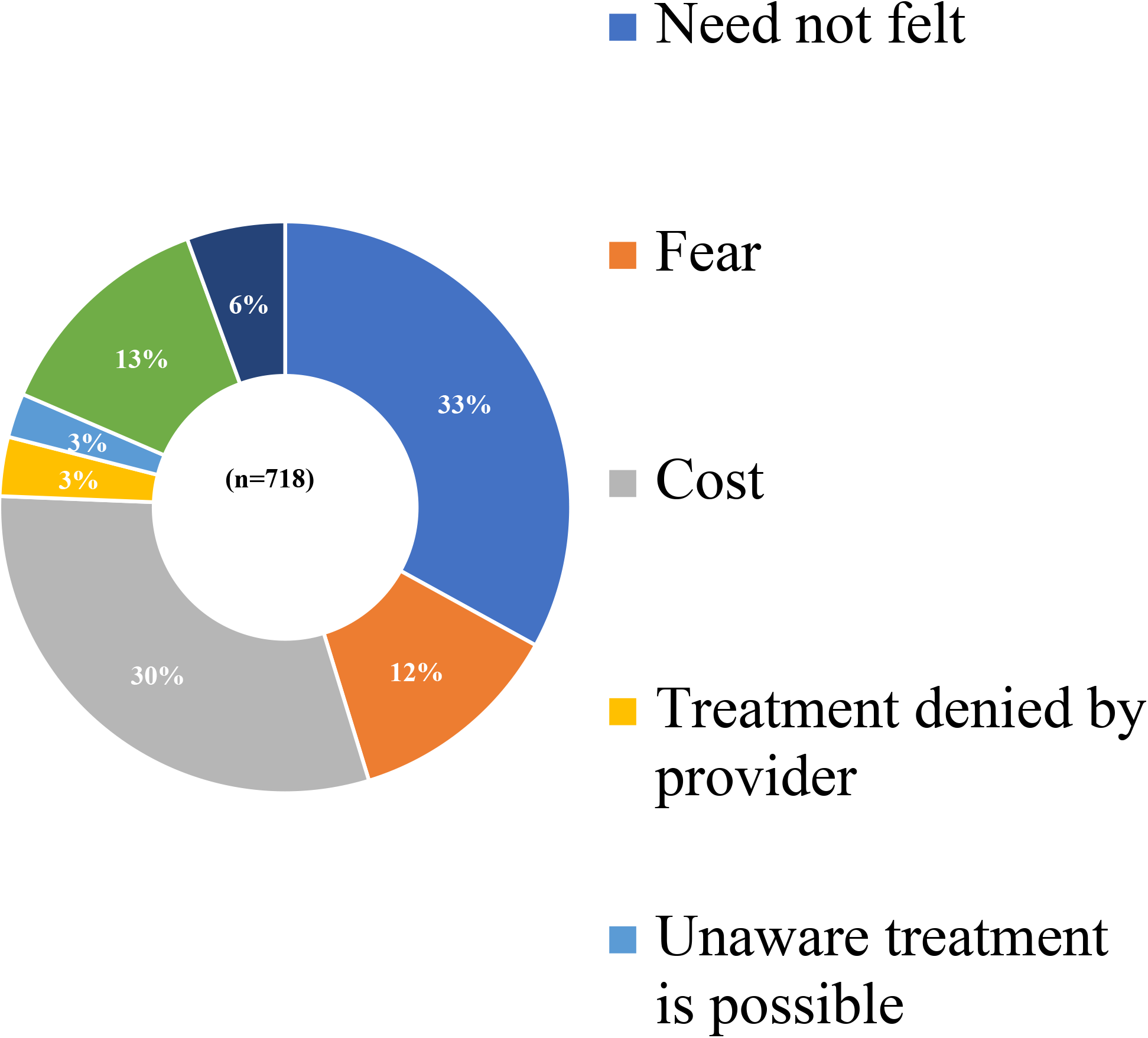

